# Deep Learning-Based Anatomical Level Classification of Craniofacial CT Slices: Comparison of CNN Architectures and Ensemble Methods

**DOI:** 10.64898/2026.10.16.26365710

**Authors:** Soh Nishimoto, Koyo Nakajima, Haruna Yoshiike, Hisako Ishise, Takuya Saito, Kenichiro Kawai

## Abstract

Computed tomography (CT) interpretation requires recognition of the anatomical level of each slice. Classification may facilitate preprocessing and AI-assisted fracture diagnosis. We developed deep learning models to classify craniofacial CT slices into six anatomical categories. A total of 11,769 images were collected from a multicenter database involving 10 Japanese university hospitals and the public Head-Neck-Radiomics-HN1 dataset. Patients were divided into training (70%) and evaluation (30%) sets. Five CNN architectures were trained from scratch, and hard- and soft-voting ensembles were evaluated. Individual-model accuracy ranged from 94.72% to 96.09%; the hard-voting ensemble achieved 96.46% accuracy (Cohen’s κ = 0.954). Although performance differed among individual models (Cochran’s Q = 99.29, p < 0.001), the ensemble did not significantly outperform EfficientNetV2B1 (McNemar’s p = 0.163). Errors occurred mainly between adjacent anatomical regions. Deep learning can classify craniofacial CT slices accurately and may support automated dataset organization and AI-assisted fracture diagnosis. External validation is needed.

## 1. Introduction

Computed tomography (CT) is widely used to diagnose and manage craniofacial trauma. It provides detailed visualization of complex anatomy and is essential for detecting facial bone fractures, assessing displacement, and planning treatment. However, interpretation requires three-dimensional anatomical knowledge and the ability to recognize subtle abnormalities across sequential two-dimensional slices. Clinicians must first identify the anatomical level of each slice before evaluating fractures, displacement, or bone defects.

Deep learning has transformed medical image analysis [1], and convolutional neural networks (CNNs) have shown promising performance in fracture detection [2]. Automated fracture diagnosis, however, presupposes that the system can identify the anatomical region represented by each image. Anatomy-specific classification has been investigated using deep CNNs [3,4], but classification of individual slices within craniofacial CT examinations has not, to our knowledge, been systematically evaluated.

Automatic anatomical classification may serve as a preliminary step in AI-based craniofacial trauma diagnosis by allowing downstream algorithms to focus on relevant regions. It may also improve the management of large imaging datasets by identifying blank images and slices from regions outside the primary field of interest. This study developed a deep learning system to classify individual craniofacial CT slices by anatomical level and compared several CNN architectures and ensemble methods.

## 2. Materials and Methods

### 2.1. CT Image Dataset

Images were obtained from two databases. The first was a multicenter database of CT examinations acquired for facial bone fracture diagnosis at 10 Japanese university hospitals: Chiba University, The Jikei University School of Medicine, Teikyo University, Kyorin University, Tokyo Women’s Medical University, Niigata University, Shinshu University, Osaka University, Hyogo Medical University, and Fukuoka University (UMIN Clinical Trials Registry ID: UMIN000039624). The study was approved by the Institutional Review Board of Hyogo Medical University (approval No. 3326). Scan conditions and slice thicknesses were not standardized. A total of 8,999 DICOM images were used.

**Figure 1.**
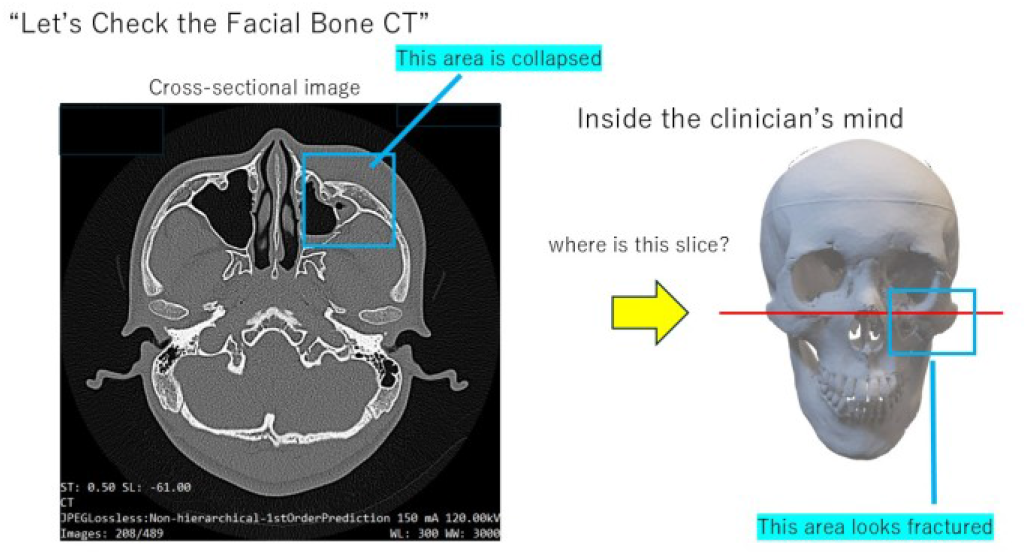
In evaluating each CT slice, the clinician would keep in mind what anatomical level the slice is.

**Figure 2.**
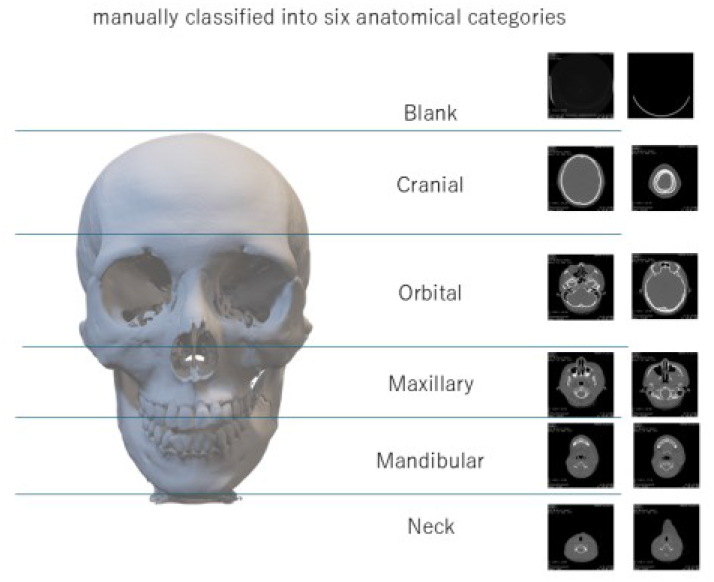
Each CT slice was manually classified into six different categories

**Table 1.** Classification performance of the five individual models and ensemble methods.

| Model | Accuracy | Balanced Accuracy | Macro Precision | Macro Recall | Macro F1 | Weighted F1 | Cohen's $\kappa$ |
| --- | --- | --- | --- | --- | --- | --- | --- |
| DenseNet | 94.72% | 94.73% | 94.11% | 94.73% | 94.40% | 94.71% | 0.9308 |
| Inception | 95.44% | 95.69% | 95.38% | 95.69% | 95.52% | 95.43% | 0.9403 |
| ResNet | 95.44% | 94.98% | 95.78% | 94.98% | 95.36% | 95.44% | 0.9402 |
| EfficientNetV2B0 | 95.44% | 95.00% | 95.77% | 95.00% | 95.37% | 95.44% | 0.9402 |
| EfficientNetV2B1 | 96.09% | 95.64% | 96.45% | 95.64% | 96.03% | 96.08% | 0.9487 |
| Hard Ensemble | 96.46% | 96.11% | 96.70% | 96.11% | 96.40% | 96.46% | 0.9536 |
| Soft Ensemble | 96.31% | 96.02% | 96.54% | 96.02% | 96.27% | 96.31% | 0.9516 |

The second dataset was Head-Neck-Radiomics-HN1, publicly available through The Cancer Imaging Archive, and comprising CT scans from patients with head and neck squamous cell carcinoma [5,6]. Axial slices at 5-mm intervals were collected, yielding 2,770 images. The combined dataset contained 11,769 images.

Patients were randomly assigned to training (70%; 7,755 images) and evaluation (30%; 4,014 images) sets at the patient level, ensuring that images from the same patient did not appear in both sets. Each slice was manually assigned to one of six categories: blank, cranial, orbital, maxillary, mandibular, or neck. Training-set counts were 178, 2,700, 1,507, 1,543, 1,112, and 715 images, respectively; evaluation-set counts were 78, 1,430, 773, 818, 653, and 262, respectively. Class imbalance was retained to reflect the distribution of real-world examinations.

### 2.2. Image Preprocessing and Data Augmentation

Each CT slice was processed as a 512 × 512-pixel two-dimensional image. During training, data augmentation included zooming (0.9–1.1), rotation (±20°), horizontal flipping, height and width shifts (0.1), and channel shift (0.15). Vertical flipping was not used because preservation of cranio-caudal orientation was essential for anatomical classification.

### 2.3. Deep Learning Models and Training

Five CNN architectures were evaluated: DenseNet121 [7], InceptionV3 [8], ResNet50 [9], EfficientNetV2B0, and EfficientNetV2B1 [10]. All models were implemented using TensorFlow-Keras applications and trained from scratch with randomly initialized weights (weights=None), without ImageNet pretraining. Training accuracy and loss, as well as validation accuracy and loss, were monitored at each epoch. The 4,014-image evaluation set was used for validation during training and for final performance evaluation.

Ensemble predictions were generated using hard voting (majority vote among predicted class labels) and soft voting (averaging predicted class probabilities) [11].

### 2.4. Statistical Analysis

Performance was assessed on the 4,014-image evaluation set using accuracy, balanced accuracy (mean recall across the six categories), macro-averaged precision, recall and F1-score, weighted F1-score, and Cohen’s kappa. A row-normalized confusion matrix was used to examine category-specific recall and misclassification patterns for the hard-voting ensemble.

Because the five models were evaluated on the same images, overall differences were tested using Cochran’s Q test. When significant, pairwise comparisons were conducted using exact McNemar tests, with p-values adjusted by the Holm method. Two-sided p < 0.05 was considered statistically significant.

## 3. Results

### 3.1. Classification Performance

The evaluation set comprised 4,014 images across six categories. The five individual models achieved accuracies ranging from 94.72% to 96.09%. Cochran’s Q test indicated a significant overall difference among models (Q = 99.29, df = 4, p < 0.001). After Holm correction, only DenseNet121 and EfficientNetV2B1 differed significantly (94.72% vs. 96.09%; adjusted p = 0.0011). Although EfficientNetV2B1 had numerically higher accuracy than InceptionV3, ResNet50, and EfficientNetV2B0, these differences were not significant after correction.

### 3.2. Individual Model versus Ensemble

The hard-voting ensemble achieved 96.46% accuracy, 0.37 percentage points higher than EfficientNetV2B1 (96.09%). Both approaches produced the same correct classification in 3,814 cases and the same incorrect classification in 99 cases. EfficientNetV2B1 correctly classified 43 images that were incorrectly classified by the hard-voting ensemble, whereas the hard-voting ensemble correctly classified 58 images that were incorrectly classified by EfficientNetV2B1. The difference in paired classification performance was not statistically significant (McNemar’s test, p = 0.163).

### 3.3. Confusion Matrix Analysis

The hard-voting ensemble achieved recalls of 97.4% for blank images, 99.7% for cranial, 96.2% for orbital, 94.1% for maxillary, 92.2% for mandibular, and 96.9% for neck images. Errors occurred mainly between adjacent regions. The most frequent error was mandibular slices classified as maxillary (7.5%); 2.8% of maxillary slices were classified as mandibular. Confusion between orbital and maxillary regions occurred in 2.3% and 2.9% of cases, respectively. In addition, 3.1% of neck images were classified as mandibular, and 2.6% of blank images as cranial. Anatomically distant categories were generally classified correctly.

## 4. Discussion

CNN models classified individual craniofacial CT slices into six anatomical categories with accuracies of 94.7–96.1%, and the hard-voting ensemble reached 96.5% accuracy with a Cohen’s kappa of 0.954. The dataset combined images from 10 hospitals with non-standardized acquisition conditions and a public head and neck cancer dataset. These results suggest that anatomical levels can be recognized reliably across heterogeneous imaging conditions, although generalizability to unseen institutions requires further assessment.

EfficientNetV2B1 achieved the highest accuracy among individual models, but after adjustment for multiple comparisons, its performance differed significantly only from DenseNet121. The hard-voting ensemble was numerically more accurate than the best individual model, but the improvement was not statistically significant. Given the computational cost of running five models, a single model such as EfficientNetV2B1 may be sufficient for practical implementation. Ensemble methods can reduce variance and improve robustness [11], and a larger evaluation dataset might clarify whether a modest benefit exists.

The error pattern was anatomically plausible. Misclassifications were concentrated at transitions between adjacent regions, particularly between the maxilla and mandible. Slices near these boundaries may contain features of both regions, including the lower maxilla, upper mandible, and teeth. Because the anatomical continuum was divided into discrete categories through manual annotation, some errors may reflect ambiguity in the labels rather than a failure of the model. For a practical workflow, a small amount of overlap between neighboring regions may be acceptable if the classification is used to narrow the region of interest for subsequent analysis rather than to establish an exact anatomical boundary.

The multicenter dataset introduced variation in scanners, acquisition conditions, and slice thickness, potentially making the task more representative of real-world data than a single-center dataset. The public dataset further increased the diversity of the images. However, the evaluation set was drawn from the same overall data sources as the training set; therefore, the effect of heterogeneity on external generalizability cannot be established. Validation using examinations from institutions and scanners not represented during development is needed.

This study has several limitations. First, the same evaluation set was used for monitoring validation performance during training and for final evaluation, which may lead to optimistic estimates. Second, labels were assigned manually, and interobserver agreement was not assessed. Third, each slice was classified independently without information from neighboring slices; incorporating sequential or three-dimensional context may reduce errors near category boundaries. Fourth, class imbalance was not corrected, and the blank category contained relatively few images, making its recall estimate less precise.

**Figure 3.**
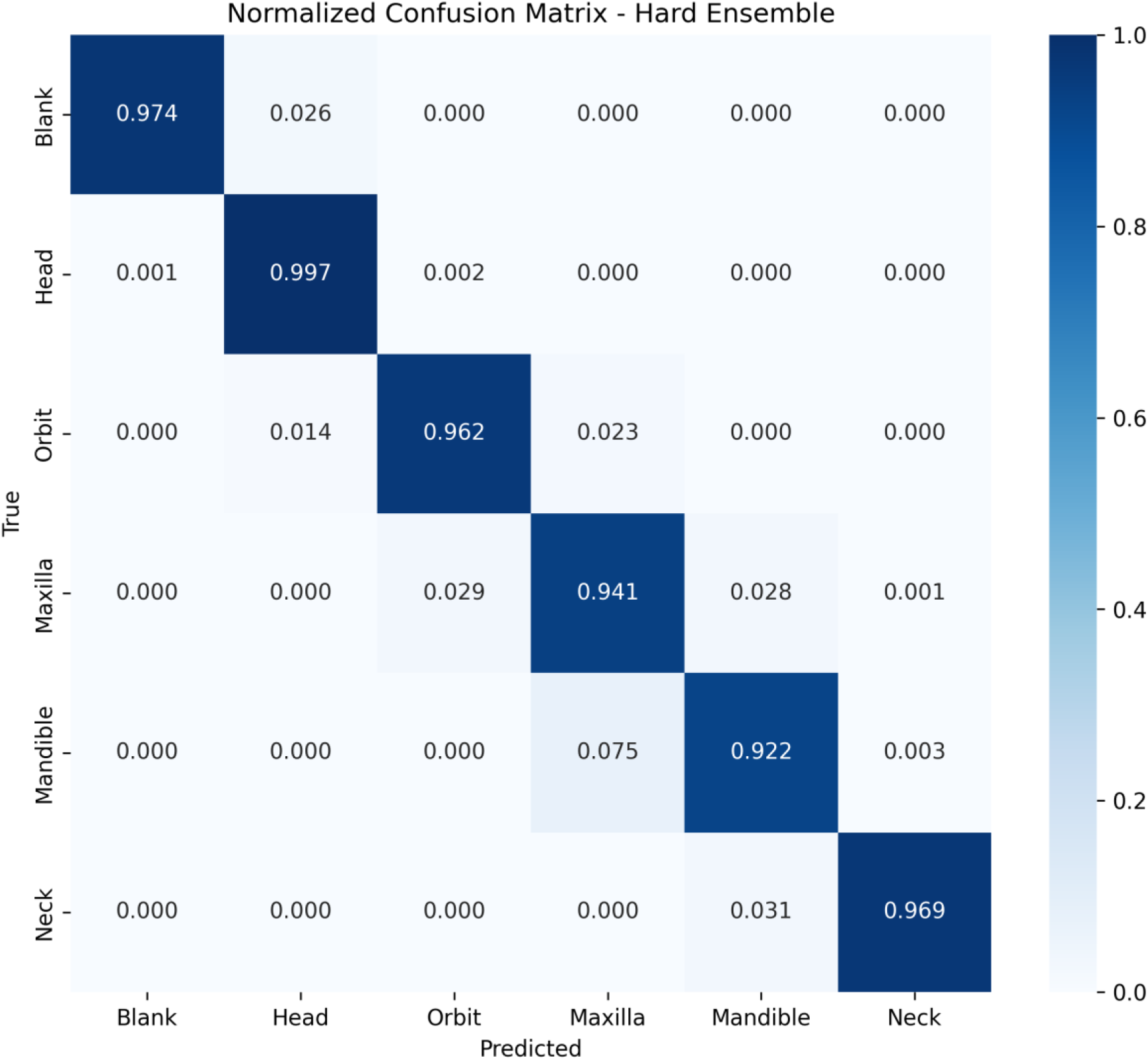
Normalized confusion matrix of the hard-voting ensemble for classification of craniofacial CT images. Rows indicate the true anatomical categories and columns indicate the predicted categories. Values represent the proportion of images within each true category assigned to each predicted category. The recall was 97.4% for blank images, 99.7% for the head region, 96.2% for the orbit, 94.1% for the maxilla, 92.2% for the mandible, and 96.9% for the neck. Misclassifications occurred predominantly between anatomically adjacent regions, particularly between the maxillary and mandibular regions.

**Table 2.** Statistical comparison of the five individual models. Overall comparison among the five models: Cochran’s Q = 99.29, df = 4, p < 0.001. Pairwise comparisons were performed using exact McNemar’s tests, with Holm correction for multiple comparisons.

| Comparison | Accuracy A | Accuracy B | Difference | Raw p | Holm-adjusted p |
| --- | --- | --- | --- | --- | --- |
| DenseNet vs Inception | 0.9472 | 0.9544 | +0.72% | 0.0215 | 0.1934 |
| DenseNet vs ResNet | 0.9472 | 0.9544 | +0.72% | 0.032 | 0.2237 |
| DenseNet vs EfficientNetV2B0 | 0.9472 | 0.9544 | +0.72% | 0.027 | 0.2162 |
| DenseNet vs EfficientNetV2B1 | 0.9472 | 0.9609 | +1.37% | <0.001 | 0.0011 |
| Inception vs ResNet | 0.9544 | 0.9544 | +0.00% | 1 | 1 |
| Inception vs EfficientNetV2B0 | 0.9544 | 0.9544 | +0.00% | 1 | 1 |
| Inception vs EfficientNetV2B1 | 0.9544 | 0.9609 | +0.65% | 0.0534 | 0.2237 |
| ResNet vs EfficientNetV2B0 | 0.9544 | 0.9544 | +0.00% | 1 | 1 |
| ResNet vs EfficientNetV2B1 | 0.9544 | 0.9609 | +0.65% | 0.0436 | 0.2237 |
| EfficientNetV2B0 vs EfficientNetV2B1 | 0.9544 | 0.9609 | +0.65% | 0.0355 | 0.2237 |

## 5. Conclusion

Deep learning models can classify craniofacial CT slices by anatomical level with high accuracy, with errors concentrated at boundaries between adjacent regions. This approach may support automated organization of CT datasets and serve as a preliminary component of AI-assisted craniofacial fracture diagnosis. External validation and further assessment of the clinical utility of region-based preprocessing are warranted.

## Data Availability

All data produced in the present study are available upon reasonable request to the authors

## Acknowledgments

We thank S. Kido, M. Hori, Y. Suzuki, H. Ishise, T. Miki, K. Yamada, T. Miyawaki, S. Moriyama, Y. Komuro, M. Fukuba, M. Ozakai, N. Oshima, K. Matsuda, Y. Sotsuka, S. Yuzuriha, T. Kubo, Y. Shikano, S. Takagi, and S. Suzuki for collecting CT data. This manuscript was partly prepared with assistance from AI-based tools (Claude, ChatGPT, Gemini, Grammarly, and GrammarCheck).

